# Supporting women who have served in the Armed Forces with a smartphone app to reduce alcohol consumption: A Randomized Controlled Trial

**DOI:** 10.64898/2026.03.22.26349029

**Authors:** Grace Williamson, Ewan Carr, Rhea Varghese, Simon Dymond, Kate King, Amos Simms, Laura Goodwin, Dominic Murphy, Daniel Leightley

## Abstract

**Background:** Alcohol misuse is common in the UK Armed Forces (AF) community, with prevalence higher than in the general population. To date, digital health initiatives to address alcohol misuse have largely focused on men, who represent around 88% of the UK AF. However, women who have served in the UK AF also drink disproportionately more than women in the general population.

**Objective:** This two-arm participant-blinded (single-blinded) confirmatory randomized controlled trial (RCT) aimed to assess the efficacy of a brief alcohol intervention (DrinksRation) compared to a web application which included NHS-focused drinking advice (BeAlcoholSmart) in reducing weekly self-reported alcohol consumption between baseline and 84-day follow-up among women who have served in the UK AF.

**Methods:** A smartphone app (DrinksRation) was compared with government guidance on alcohol use. The app included features tailored to the needs of women who have served and was designed to enhance motivation to reduce alcohol consumption. The trial enrolled women who had completed at least one day of paid service in the UK Armed Forces. Recruitment, consent, and data collection were completed automatically through the platform. The primary outcome was the between-group difference in change in self-reported weekly alcohol consumption from baseline to day 84, measured using the Timeline Follow-Back method. The secondary outcome was the between-group difference in change in Alcohol Use Disorders Identification Test (AUDIT) score from baseline to day 84. Process evaluation outcomes included app engagement and usability, with usability assessed using the mHealth App Usability Questionnaire.

**Results:** A total of 88 women UK AF veterans were included in the final analysis (control=37; intervention=51). At 84 days post-baseline, participants in the intervention group (DrinksRation) showed a greater reduction in weekly alcohol consumption compared to controls (BeAlcoholSmart) (adjusted mean difference in change from baseline = −11.6 units; 95% CI: −19.7 to −3.6; *p*=0.005). AUDIT scores decreased more in the intervention group (adjusted mean difference in change = −3.9; 95% CI: −6.9 to −1.0; *p*=0.01). Usability scores at day 28 were significantly higher for the intervention group across all domains. No serious adverse events or technical issues were reported.

**Conclusions:** DrinksRation reduced alcohol consumption and hazardous drinking among women who have served in the UK Armed Forces. Engagement was strong, usability was high, and no safety concerns were identified. These findings support the potential of tailored digital interventions to address alcohol misuse in women who have served in the UK Armed Forces.

**Registration:** ClinicalTrials.gov (trial registration: NCT05970484).

## Introduction

There are an estimated 1.8 million Armed Forces (AF) veterans in England and Wales, defined by the UK Government as anyone who has served in the military for at least one paid day. Around 12% are estimated to identify as women/female^1^. Women have served in the AF for more than a century, and their contributions have been recognized by successive UK governments and the wider public^2,3^. However, concerns remain about deep seated military culture and the ways in which evolving Service requirements may affect the health and wellbeing of many who have served ^2,3^. This also includes limited evidence on the impact of alcohol use on women’s health^4,5^.

Evidence on alcohol use among female veterans is limited, but what has been published has indicated a concerning pattern of increasing consumption and elevated risk^6^. The limited existing evidence suggests that alcohol use among women who served is increasing, and that they are significantly more likely to report hazardous drinking compared with their civilian counterparts^7^. In the UK, hazardous alcohol consumption is commonly defined as consuming more than 14 units of alcohol per week on a regular basis, meaning a level of drinking that increases the risk of harm. Palmer and colleagues (2021) reported high levels of hazardous drinking, finding that almost half of female veterans (49%; 389/779) were drinking at a hazardous level or higher, which is considered harmful to health^8^. The UK Chief Medical Officers recommend that adults (both male and female) limit intake to no more than 14 units per week to reduce the health risks associated with alcohol consumption^9^.

Alcohol misuse often co-occurs with common mental health disorders, including PTSD, anxiety, or depression, and alcohol is frequently used as a coping mechanism^2,10,11^. Common mental health disorders are more common in female than male veterans^7^. Evidence suggests that, like male veterans, female veterans face barriers to accessing mental health support, with alcohol misuse contributing to delayed help-seeking and engagement with care^12,13^. Although female veterans typically drink less than male veterans, their rates of hazardous drinking are higher than women in the general population, placing them at increased risk of poorer health outcomes^14^.

In recent years, there has been an increasing unmet need in treatment for mental health in the UK, with patients waiting longer for treatment for alcohol use. To help address this, we developed DrinksRation^15^, a digital intervention designed to support the AF Community, serving^16^ and veterans, in managing and reducing the amount they drink^17–20^. DrinksRation is unique in that the app content is tailored to the AF; messaging is dynamic and tailored using behavior change techniques (BCT) to promote positive changes in lifestyle choices.

DrinksRation is the only app targeting alcohol use in the UK AF veteran community. It is designed to (1) overcome geographical limitations; (2) use wearable technology to inform decision-making and personalization; (3) avoid the stigma associated with receiving help in person; and (4) provide convenience since users can use the app as they prefer (discretely or openly). The app is freely available via Apple App store. DrinksRation is supported by a robust evidence base, including a randomized controlled trial that demonstrated that the app is efficacious in reducing alcohol consumption in the AF veteran population^18^.

DrinksRation was initially developed to support veterans and did not explore the nuances of gender, nor acknowledge people’s differing motivations to consume alcohol. A review of DrinksRation highlighted a need for feminist intersectionality in digital health to incorporate the unique needs of females^21,22^. Digital health technologies can bolster gender equality through increased access to healthcare, empowerment of one’s health data, overcoming the specific barriers facing female veterans, and reducing the burden on healthcare systems^23– 25^. In this study, we conducted a randomized controlled trial (RCT) to assess the efficacy DrinksRation in reducing self-reported weekly alcohol consumption by day 84 follow-up among female veterans in who drink at hazardous or harmful levels.

## Methods

### Ethics

This study was approved by the ethics committee of King’s College London (LRS/DP-22/23-36879). This study was prospectively registered on ClinicalTrials.gov (trial registration: NCT05970484). The trial protocol was also published prior to the collection of any data; see Williamson and colleagues (2023^26^) for more information.

### Trial design

This was a two-arm, participant-blinded (single-blinded) confirmatory RCT, comparing (intervention arm) the DrinksRation smartphone app with (control arm) a progressive web application (PWA) presenting NHS-focused drinking advice (named BeAlcoholSmart; BAS). DrinksRation provided individualized normative advice with features designed to enhance participant motivation, interactive feedback, self-efficacy in modifying their alcohol consumption, and personalized gender-specific messaging. We hypothesized that the intervention arm would be efficacious in reducing alcohol consumption compared with the control arm. Participants in both arms were asked to use the digital interventions for 12 weeks (84 days).

### Procedure and participants

Participants were recruited between January 2024 and July 2024 through networking and social media following published recruitment strategies^27–29^. Throughout the recruitment period, emails were periodically sent to a curated list of 336 organizations that supported the Armed Forces Community in the UK AF. These organizations represented voluntary, community, health and commercial sectors, with all having some focus on veterans’ health. Organizations shared recruitment materials via newsletters, social media, printed flyers at events and shared with internal networks. We also recruited participants via paid promotional advertisements on social media (e.g. X/Twitter, Facebook, Instagram, and LinkedIn) with a link enabling potential participants to complete the eligibility and consent survey.

Potential participants who responded to the email invitation or social media adverts were provided a link to the study website, which contained an explanation of the study, a link to the participant information sheet, and an invitation to complete the eligibility and consent survey. If participants met the eligibility criteria, they were randomized and sent instructions on accessing the BeAlcoholSmart or DrinksRation app using a unique quick response (QR) code. Once they had accessed the platform, they could then register an account and complete the baseline questionnaire (day 0). If a participant did not complete signup, a maximum of three follow-up attempts were made to support enrolment.

Participants were included in the study if they: were aged 18 years or older; identified as female (self-reported gender); lived in the UK; consumed more than 14 UK units (approximately 120g of pure ethanol) of alcohol or more per week as measured using Timeline Follow-back for Alcohol Consumption (TLFB^30^) at baseline (day 0); were veterans of the UK AF, defined as having completed at least one day of paid employment in the UK AF (via self-report at eligibility screening); and owned a iOS or Android smartphone.

### Randomization and blinding

Participants were randomized (1:1) to either DrinksRation or BeAlcoholSmart. Randomization was conducted automatically when a token code was generated alongside a unique proxy identifier (managed by GW). At this point, participants were automatically randomized to the control or intervention arm and were blinded. The randomization procedure was carried out automatically via a randomization algorithm with no human involvement except to provide a proxy identifier. Since allocations were generated dynamically, it was not possible to view future allocations. DL and GW were unblinded to treatment allocation to enable the management of DrinksRation or BeAlcoholSmart, participant recruitment and retention. EC was unblinded to treatment allocation to prepare statistical reports at the end of data collection. DL and EC were not involved in any participant engagement. All other members of the study team were blind to treatment allocation.

### Sample size

A power calculation was performed based on previously reported data from the DrinksRation RCT^18^. With 148 participants (74 per arm), we will have at least 80% power to detect a difference in alcohol consumption of three units (approximately 24g of pure ethanol per week) between the intervention and control arm at the 84-day follow-up assessment. This calculation assumes a standard deviation of 5.6 units, attrition of 40% by day 84, a correlation of 0.7 between successive follow-up assessments, and an alpha of 5%.

We chose a minimum difference of three units based on treatment effects observed in similar studies^31–33^ and reductions observed in the DrinksRation feasibility trial^17^ and confirmatory RCT in help-seeking veterans^18^, which found reductions at 28 days of 22 units between the arms. We have chosen a smaller minimum difference of 3 units, recognizing that treatment effects for brief interventions reduce over time, and the target population for this study is not a help-seeking population. This was also the average unit decrease observed in a review of digital brief alcohol interventions by Kaner and colleagues (2018^34^).

### Interventions

The DrinksRation (formerly called *InDEx*^19,20,35^) app was developed following the Medical Research Council Complex Intervention Guidelines^36^ and using a co-design methodology. It was developed by researchers from King’s Centre for Military Health Research (King’s College London) and Lancaster University, supported by experts in smartphone app development, epidemiology, addiction psychiatry, and military mental health. The app was designed as a Research Viable Product^37^ to support veterans drinking at a hazardous or harmful level by providing bespoke advice and support.

The app seeks to enhance participants’ motivation and self-efficacy in modifying their alcohol consumption using BCTs which included in-app content (see **Error! Reference source not found**.) and push notifications. The iterative development process, theoretical framework, feasibility trial, pilot, and RCT are reported elsewhere^18–20,35,38^. Briefly, DrinksRation was developed and tested with five core modules (see Figure 1 for example screenshots), these are: account management, questionnaire and individualized normative feedback, self-monitoring and feedback, goals (setting and review) and personalized messaging. In this study, DrinksRation was modified to cater to the specific needs of women who have served, this includes specific content, messaging, alcoholic drinks and the ability to further customize the presentation of a range of metrics.

**Figure 1.**
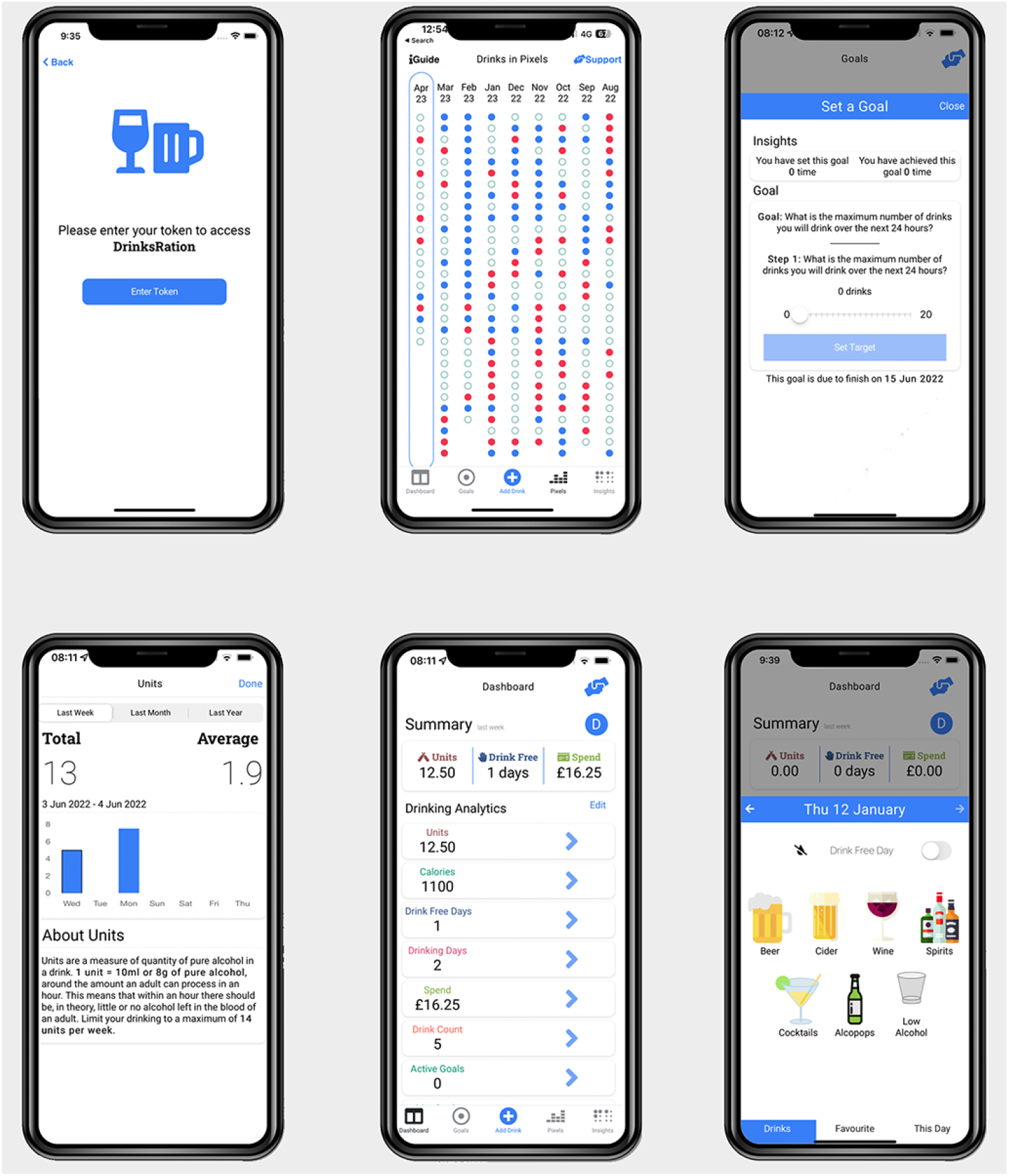
Example screenshots extracted from the DrinksRation app.

Participants in the intervention arm were asked to complete all questionnaires via the app, including additional weekly questionnaires on their mood and general mental health, which were used to personalize the app content and push notifications.

The control arm received access to BeAlcoholSmart. BeAlcoholSmart was deployed as a PWA. A PWA is a website that looks and behaves like a smartphone app. The PWA takes advantage of smartphone features such as push notifications and haptics without requiring the participant to download via an app store. The BeAlcoholSmart was accessible to participants in the control arm via a unique link. The PWA contained a 7-day alcohol unit calculator and generic public health guidance on safe drinking (see for further information^39^). Participants received reminders via email prompting them to consult the guidance as part of the BeAlcoholSmart. Control participants were invited to complete all questionnaires via Qualtrics, with an email reminder being sent when they were due.

### Incentivization

After completing outcome questionnaires (day 84) at the primary endpoint (day 84), participants received a £20 Love2Shop voucher. This was used to encourage completion of the outcome measure. No cash alternative was offered.

### Messaging

Participants in the intervention arm (DrinksRation) received personalized push notifications to prompt them to use the drinks diary, engage with the app and complete outcome measures, suggest alternative behaviors, provide feedback on goals, and promote a healthy lifestyle. We used a bank of 180 messages developed previously (see ^19^ for a detailed description), informed by the Health Action Process Approach framework and targeting specific BCTs.

DrinksRation used baseline and weekly questionnaires to inform participant messaging and provide an individualized participant-centric approach. Baseline measurements were used to identify suitable messages, and as a participant engaged with DrinksRation, regular measurements, including questionnaires (baseline and weekly questionnaires) and the drinks diary, were used to reflect current behaviour and attitudes. The notifications covered a range of topics to target beliefs and motivations with the primary aim of increasing the participants’ awareness of their drinking habits and behaviors. The notifications were divided into three categories:

1. Tailored: personalized to drinking habits, baseline, and weekly questionnaires;
2. Tailored and triggered: tailored to baseline and continuous measurements and a specific event occurring; and
3. Targeted (generic): sent on specific days to highlight inactivity, reminders to complete a questionnaire, or to alert participants to a new feature of the app.

The DrinksRation messaging framework decided when a message should be sent based on user analytics data.

To increase engagement and maximize completion of the follow-up measures, participants using DrinksRation were also sent reminder emails one day before a monthly questionnaire was due.

For the control arm (BeAlcoholSmart), participants only received notifications (via email) when they joined the platform and when outcome measures were due. The welcome email contained information on how to use the BeAlcoholSmart platform and when future notifications would be sent. The questionnaire emails included a reminder to consult the BeAlcoholSmart platform and complete the questionnaire due at this time point. No other notifications were sent.

### Consent and withdrawal

Informed consent from all participants was obtained via the eligibility and consent survey before the collection of any personal data. For participants in the intervention arm, we asked for specific on-device permissions to enable the full functionality of the DrinksRation app. For example, to receive push notifications or share health tracking data. Participants could change these optional permissions at any time via the ‘settings’ page of the app.

Participants were informed via the participant information sheet that they could withdraw from the study or withdraw from data collection at any time. For participants who withdrew from the study: (i) we attempted to collect follow-up information if they agreed to continued data collection; (ii) we retained their existing data if they agreed to this use. Alternatively, withdrawn participants could delete their study information by contacting the research team. For those in the control arm, they needed to withdraw by contacting the research team. For those in the intervention arm, they could withdraw via the app.

If participants withdrew, they were asked to delete the intervention app and were denied further access. Suspected fake or bot participants were reviewed and flagged for consideration during the analysis (see ^40^ for further information).

### Study measures

Study measures and data collection timepoints in this study are detailed in Table 1.

**Table 1:** Summary of measures and data collection timepoints (day 0, 7, 12, 21, 28, 56 and 84).

| Day/Measure | Days post-randomization |  |  |  |  |  |  |
| --- | --- | --- | --- | --- | --- | --- | --- |
|  | 0<br>(Baseline) | 7 | 14 | 21 | 28 | 56 | 84<br>(Primary endpoint) |
| **Questionnaires** |  |  |  |  |  |  |  |
| Informed consent(s) | I/C |  |  |  |  |  |  |
| Socio-demographic | I/C |  |  |  |  |  |  |
| Depression (PHQ2) <sup>41</sup> | I/C | I | I | I | I | I/C | I/C |
| Anxiety (GAD2) <sup>42</sup> | I/C | I | I | I | I | I/C | I/C |
| International Trauma Questionnaire for PTSD (ITQ-PTSD; <sup>43</sup>) | I/C |  |  |  | I/C | I/C | I/C |
| Readiness to Change Ruler <sup>44</sup> | I/C |  |  |  | I/C | I/C | I/C |
| Self-efficacy Ruler <sup>44</sup> | I/C |  |  |  | I/C | I/C | I/C |
| Alcohol Use Disorder Identification Test (AUDIT) <sup>45</sup> | I/C |  |  |  | I/C | I/C | I/C |
| EQ-5D-5L <sup>46</sup> | I/C |  |  |  | I/C | I/C | I/C |
| 7-day Timeline Follow Back for Alcohol Consumption (TLFB; <sup>30</sup>) | I/C |  |  |  | I/C | I/C | I/C |
| Adverse event reporting |  |  |  |  | I/C | I/C | I/C |
| **Usability evaluation** |  |  |  |  |  |  |  |
| mHealth App Usability Questionnaire (MAUQ) <sup>47</sup> |  |  |  |  | I/C | I/C | I/C |
| **Remote data collection** |  |  |  |  |  |  |  |
| Wearable sensors* | Continuous: I/C |  |  |  |  |  |  |
| Smartphone sensors* | Continuous: I/C |  |  |  |  |  |  |

I: Intervention Arm data collected via app; C: Control Arm data collected via Qualtrics. \*Additional participant consent is required.

**Table 2:** Participant characteristics by allocation.

| Characteristic | Overall (n=88) | BAS (n=37) | DrinksRation (n=51) |
| --- | --- | --- | --- |
| Age at enrolment (years; SD) | 47.38 (9.96) | 47.18 (10.58) | 47.52 (9.58) |
| Gender |  |  |  |
| Female | 88 (100) | 37 (100) | 51 (100) |
| Relationship status |  |  |  |
| Married | 38 (43.18) | 17 (45.95) | 21 (41.18) |
| Living with partner | 20 (22.73) | 10 (27.03) | 10 (19.61) |
| In long term relationship | 10 (11.36) | 3 (8.11) | 7 (13.73) |
| Single | 9 (10.23) | 2 (5.41) | 7 (13.73) |
| Separated | 5 (5.68) | 2 (5.41) | 3 (5.88) |
| Divorced | 5 (5.68) | 2 (5.41) | 3 (5.88) |
| Widowed | 1 (1.14) | 1 (2.70) | 0 |
| Employment status |  |  |  |
| Working part-time | 19 (21.59) | 8 (21.62) | 11 (21.57) |
| Working full-time | 44 (50.0) | 18 (48.65) | 26 (50.98) |
| Unemployed | 7 (7.95) | 3 (8.11) | 4 (7.84) |
| Economically inactive | 4 (4.55) | 3 (8.11) | 1 (1.96) |
| Retired | 8 (9.09) | 3 (8.11) | 5 (9.80) |
| Education/training | 4 (4.55) | 0 | 4 (7.84) |
| Not reported | 2 (2.27) | 3 (8.11) | 0 |
| Military branch |  |  |  |
| Royal Navy | 21 (23.86) | 9 (24.32) | 12 (23.53) |
| Army | 48 (54.55) | 21 (56.76) | 27 (52.94) |
| Royal Air Force | 15 (17.05) | 6 (16.22) | 9 (17.65) |
| Other* | 4 (4.55) | 1 (2.70) | 3 (5.88) |
| Service length (years; SD) | 12.39 (7.61) | 11.83 (6.30) | 12.81 (8.50) |
| Probable PTSD caseness | 27 (31.03) | 16 (43.24) | 11 (22.00) |
| Probable Depression caseness | 46 (52.27) | 21 (56.76) | 25 (49.02) |
| Probable Anxiety Caseness | 34 (38.64) | 13 (35.14) | 21 (41.18) |
| AUDIT Score (mean; SD) | 14.47 (6.91) | 15.13 (7.37) | 14 (6.60) |
| AUDIT Caseness |  |  |  |
| Low risk ($\leq 7$) | 16 (18.18) | 5 (13.51) | 11 (21.57) |
| Harmful/hazardous ($8 \leq \leq 15$) | 30 (34.09) | 13 (35.14) | 17 (33.33) |
| High risk/problematic ($\geq 16$) | 42 (47.73) | 19 (51.35) | 23 (45.10) |
| Baseline alcohol weekly units (mean; SD) | 27.23 (16.10) | 28.28 (17.44) | 26.47 (15.18) |

\*Other types including tri-service roles.

### Baseline measures

After downloading the respective platform, participants were asked to complete a baseline questionnaire to assess physical and mental health, readiness to change, self-efficacy, health status and sociodemographic factors (e.g. age, gender, ethnicity, employment status, and occupation). The questionnaire schedule is presented in Table 1.

### Primary outcome

The primary outcome in this study was self-reported alcohol consumption measured by the 7-day TLFB. At baseline (day 0) and 28-, 56-, and 84-days post-randomization, participants were asked to report the number and type of drinks that they consumed in each of the previous seven days via DrinksRation or BeAlcoholSmart. Using standard unit measurements (see **Error! Reference source not found**.), we calculated weekly alcohol consumption for baseline and each follow-up by summing the number of units assigned to each drink. To maximize completion of the primary outcome at each follow-up, participants were asked to complete the primary outcome measure before completing the rest of the questionnaire.

### Secondary outcome

The secondary outcome is the change in AUDIT score, measured at baseline (day 0) and day 84 follow-up between the control and intervention groups.

#### Usability

We examined the usability of the DrinksRation and BeAlcoholSmart using mHealth App Usability Questionnaire (MAUQ^47^) at day 28.

### Safety information

Adverse Events and Serious Adverse Events were monitored throughout the intervention period via (1) direct contact from participants to GW; and (2) a questionnaire presented to participants via the app on days 28, 54, and 84 post-randomization, asking questions about risk. All reported events were reviewed by a clinician (researcher DM), who, where required, contacted participants to perform a clinical interview and risk assessment.

### Fraudulent and suspicious participants

Potentially fraudulent or suspicious participants were identified using several methods embedded within the survey and app management systems^40^. Specifically, these methods included analysis of IP address patterns, email address validation (such as invalid or disposable email domain), absence of email verification, and internal technical validation checks within the DrinksRation and BeAlcoholSmart platforms. Where suspicion arose, confirmation of veteran status was sought through manual verification procedures. Two investigators (GW and SD) independently reviewed flagged participant details to confirm suspicion and determine participant exclusion. Participants confirmed as suspicious or fraudulent were subsequently excluded from further participation and analyses.

### Statistical analyses

Participant characteristics were summarized by trial arm using descriptive statistics: frequencies and percentages for categorical variables and means with standard deviations (SD) for continuous variables.

The primary outcome (weekly alcohol consumption) and secondary outcome (AUDIT score) were analyzed using mixed models for repeated measures. Each model included baseline outcome value, treatment allocation (0=control, 1=intervention), time since baseline (days 28, 56, and 84), a treatment-by-time interaction, and age as a covariate. Repeated measures within participants were modelled using an unstructured covariance matrix.

Degrees of freedom and standard errors were adjusted using the Kenward-Roger method ^48^.

Treatment effects were estimated as adjusted mean differences between arms at each follow-up time point, with 95% confidence intervals (CIs). The primary endpoint was day 84. As a descriptive measure of effect size, a standardised mean difference was calculated by dividing the adjusted mean difference at day 84 by the pooled observed standard deviation at that time point. Statistical significance was defined as *p* < 0.05.

Analyses were conducted on a modified intention-to-treat basis, including all randomized participants except those identified post-as automated or duplicate registrations (suspected bots) according to a priori criteria described elsewhere. Participants were required to have at least one post-baseline outcome measurement to be included in the outcome analyses.

Missing outcome data were handled implicitly within the primary mixed models for repeated measures using likelihood-based estimation under a missing-at-random (MAR) assumption, conditional on variables included in the model. All analyses were conducted using Stata or R (v4.5.2 ^49^). Mixed models for repeated measures were fitted in R using *mmrm* (v0.3.17; ^50^) and *emmeans* (v2.0.1^51^).

#### Process evaluation

We examined process evaluation measures as a proxy for app usage. These were reported as median and interquartile range. These are reported in four categories: (1) app utilization based on app analytics data provided by Google Analytics (California, USA), (2) drinking analytics based on server interactions, (3) weeks active based on a participant having a minimum of 3 server interactions in a calendar week; and (4) notifications sent by the server.

#### Usability

We examined usability (MAUQ) at day 28. Questionnaire responses were aggregated into (1) overall usability, (2) ease of use, (3) interface and satisfaction, and (4) usefulness. Results were summarized with means and SDs.

#### Reporting

The study was reported following the Template for Intervention Description and Replication^52^ and CONSORT (Consolidated Standards of Reporting Trials^53^; the e-Health version^54^) checklist.

## Results

### Study participation, sample characteristics, and attrition

Participants were recruited through advertisements on social media and research databases (see Figure 2). A total of 1,247 individuals consented and completed eligibility screening. Of these, 987 were excluded due to ineligibility criteria, the majority due to not meeting the 14-unit threshold for alcohol consumption (*n*=427; 43%) or incomplete responses (*n*=313; 32%). Additionally, 36 participants were excluded due to suspected fraudulent activity or automated scripting. Therefore, 260 eligible participants were randomized and invited to access the intervention or control. Of these, 150 created an account; 59/150 (39.33%) of those invited to the control arm (BAS), 91/150 (60.67%) of those invited to the intervention (DrinksRation).

**Figure 2.**
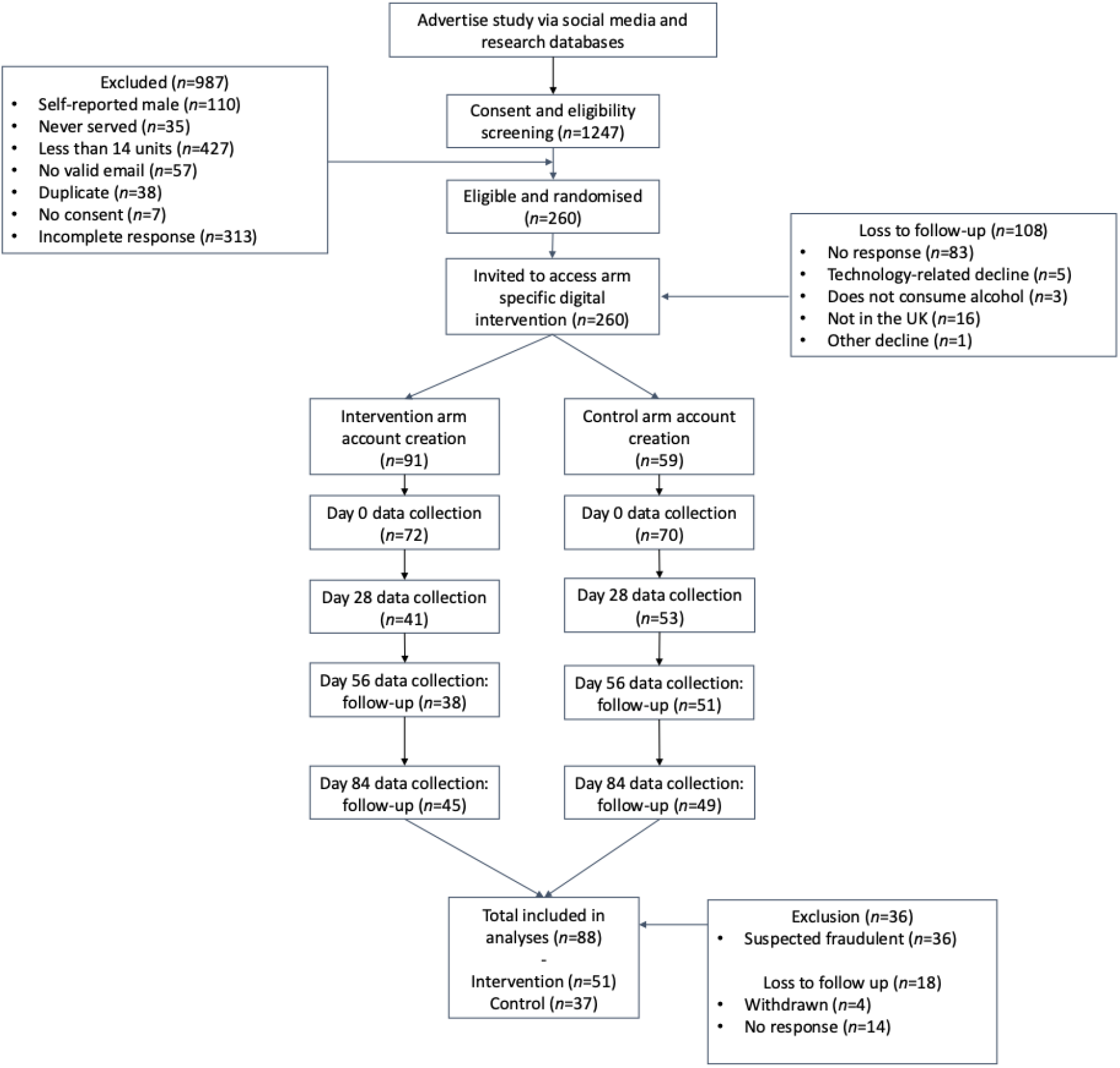
CONSORT diagram of the study flow and allocation.

A total of 88 female participants were included in the final analysis (BAS *n*=37, DrinksRation *n*=51), with an overall mean age of 47.5 years (SD=9.98). Participants were predominantly married or living with a partner (65%), employed full-time (50%), and had served primarily in the Army (55%). The mean length of military service was approximately 12 years (SD=7.58). At baseline, nearly half of participants reported probable depression (51%), over a third had probable anxiety (39%), and almost a third met criteria for probable PTSD (31%). The mean AUDIT score was 14.25 (SD=7.06), indicating hazardous drinking patterns across the sample, with participants consuming an average of 27 units of alcohol per week.

### Primary outcome

At baseline, mean weekly alcohol consumption was 28.3 units (SD = 17.4) in the control group (BAS) and 26.5 units (SD = 15.2) in the intervention group (DrinksRation) (Table 3). At 84 days, participants in the DrinksRation group showed a greater reduction in alcohol use than those in BAS. The adjusted mean difference in change from baseline was −11.6 units/week (95% CI: −19.7 to −3.6; *p*=0.005), indicating a statistically significant effect in favour of the intervention. The corresponding standardised effect size at day 84 was 0.67 (95% CI 0.16 to 1.20), based on the pooled observed standard deviation at that follow-up, consistent with a medium-to-large effect.

**Table 3:**
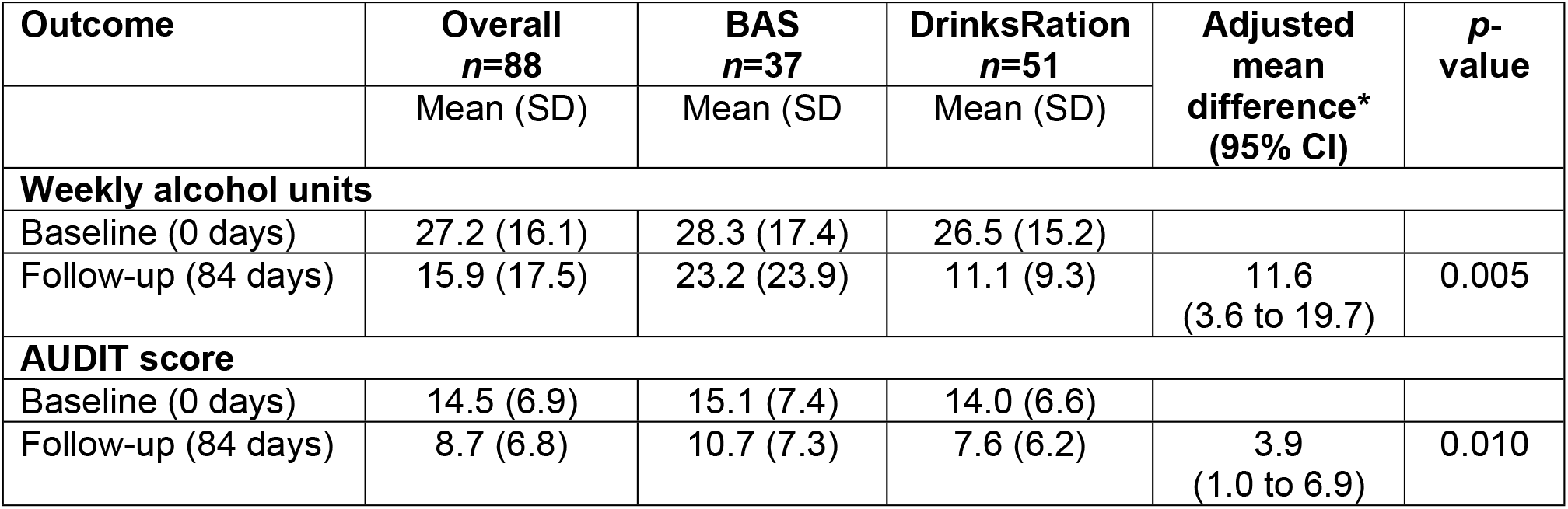
Analysis of change in weekly alcohol consumption and AUDIT scores from baseline to day 84.

### Secondary outcome

Table 3 also shows changes in AUDIT scores from baseline to 84 days. At baseline, mean AUDIT scores were 15.1 (SD = 7.4) in the BAS group and 14.0 (SD = 6.6) in the DrinksRation group. At follow-up, scores had decreased to 10.7 (SD = 7.3) in BAS and 7.6 (SD = 6.2) in DrinksRation. Participants in the DrinksRation arm showed a greater reduction in AUDIT scores over time than those in BAS. The adjusted mean difference in change from baseline was −3.9 points (95% CI: −6.9 to −1.0; *p*=0.010), favouring the intervention. The corresponding standardised effect size at day 84 was 0.58 (95% CI 0.11 to 1.10), based on the pooled observed standard deviation at that follow-up, consistent with a medium effect.

These findings indicate a clinically meaningful reduction in alcohol consumption and related harm among participants.

### Process evaluation

Participants in the intervention group (DrinksRation) exhibited greater engagement compared to the control group (BAS) Table 4. Specifically, intervention participants initiated the app more frequently (median initializations: intervention = 13, control = 1), engaged in a higher number of sessions (median session count: intervention = 132.5, control = 2), and maintained engagement over a longer period (median weeks active: intervention = 11, control = 1). However, the median duration of individual sessions was shorter in the intervention arm compared to the control arm (17.45 seconds vs. 33.86 seconds, respectively). Intervention participants logged a median of 19 (IQR: 5 to 22) drinking days and 19 (IQR: 5 to 51) drink-free days, consuming approximately 5.86 units per drinking day. Push notifications (median = 52 notifications per participant) were frequently delivered to the intervention group, while participants in the control arm received a median of 7 email notifications over the study period.

**Table 4:** Engagement and app interactions over the study period per participant stratified by allocation.

| Interactions | BAS<br>median (IQR) | DrinksRation<br>median (IQR) |
| --- | --- | --- |
| **Engagement measures** |  |  |
| Initializations | 1 (1 to 3) | 13 (7 to 39) |
| Session count | 2 (1 to 3) | 132.5 (54 to 284) |
| Session duration (seconds) | 33.86 (18.87 to 55.50) | 17.45 (10.44 to 47.95) |

| **App-recorded interactions** |  |  |
| --- | --- | --- |
| Drinking days | N/A* | 11 (4 to 22) |
| Drink-free days | N/A* | 19 (5 to 51) |
| Units consumed per drinking day | N/A* | 5.36 (4.44 to 7.11) |
| **Notifications** |  |  |
| Push notifications | - | 52 (33 to 67) |
| Email | 7 (5 to 9) | - |
| Weeks active | 1 (1 to 2) | 11 (8 to 15) |

\*not applicable; participants in the control arm were not able to provide this information.

#### Usability

Table 5 demonstrates the usability ratings stratified by allocation, with higher scores in the intervention arm compared with the control arm. Participants receiving DrinksRation reported higher ease-of-use (mean = 6.06 v 5.51), greater satisfaction with the app interface (mean = 6.29 v 5.18), and higher perceived usefulness (mean = 6.28 v 4.93). Overall usability was also rated significantly higher in the intervention group (mean = 6.22) compared to the BAS (mean = 5.22).

**Table 5:**
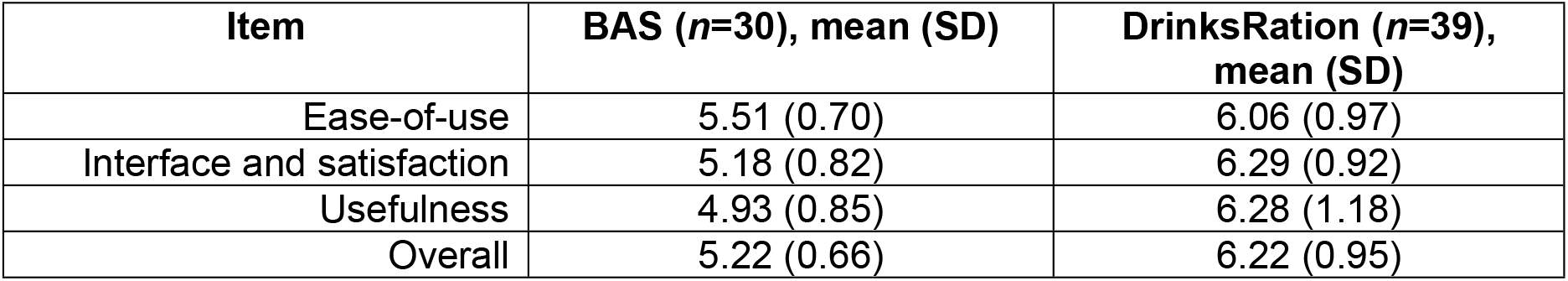
mobile health (mHealth) App Usability Questionnaire results at day 28, stratified by allocation.

### Adverse events and technical issues

Table 6 outlines self-reported adverse events. Overall, adverse events reported during the study were infrequent and generally minor in both arms, with no serious safety concerns or life-threatening incidents identified. No technical issues were reported with the DrinksRation app.

**Table 6:**
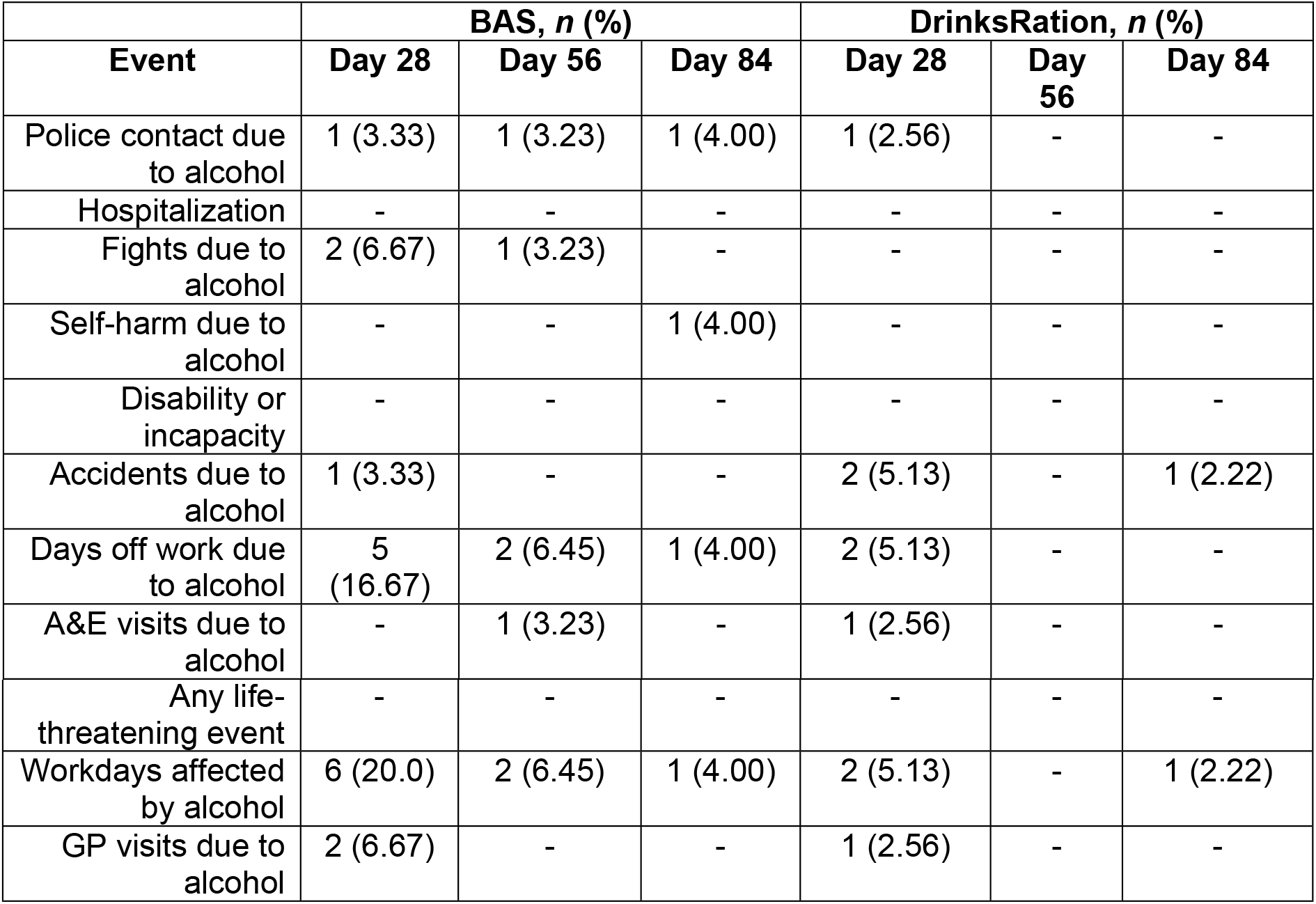
Self-reported adverse events across the study period. All questions were framed because of alcohol consumption on the prior 28 days.

## Discussion

This RCT evaluated the efficacy of DrinksRation, tailored specifically for female UK AF veterans in the UK AF consuming alcohol at hazardous or harmful levels. Our findings demonstrate significant reductions in weekly alcohol consumption and AUDIT scores at 84 days post-randomization among participants using DrinksRation compared with the control arm. This indicates both statistical significance (via alcohol unit decrease) and clinical relevance (AUDIT score decrease). These results align with previous research that supports the effectiveness of personalized digital interventions for reducing hazardous alcohol consumption^55,56^.

Participants in this study had a mean reduction of 8.7 units at day 84, and a fall in AUDIT score of 5.4 points. For context, an earlier RCT for DrinksRation^55^ in help-seeking, predominantly male, veterans obtained a 28.2 unit reduction from a baseline of 55 units. Broadly, Kaner and colleagues (2017)^34^ review of digital alcohol brief interventions reported an average decrease of 23g ethanol (3 UK units) per week from a mean baseline of 244g (30.5 units) in community drinkers. We believe that while the baseline alcohol use in our sample (27 units) is lower than the Kaner and colleagues (2017)^34^ average, the observed reduction is almost three times larger. This outcome is unlikely to be explained solely by a higher starting point. Instead, it may indicate DrinksRation tailoring (e.g. gender-sensitive messaging, military-relevant terms, personalized content) may deliver an effect size substantially above that seen with generic digital interventions.

Similar outcomes have been documented in other digital interventions targeting alcohol misuse, underscoring the importance of tailored app content and behavioural prompts in achieving sustained behavioural changes^56–58^.These findings suggest that personalized feedback and motivational messaging, core components of DrinksRation, may have contributed to self-efficacy and motivation, aligning with behaviour change theories such as the Health Action Process Approach^59^.

Engagement metrics indicated active and sustained use within the intervention group, consistent with prior literature emphasizing the role of personalized push notifications in maintaining and promoting user engagement^60^. Participants assigned to DrinksRation demonstrated significantly higher app interaction rates, session counts, and overall engagement compared to those in the control arm, despite shorter individual session durations. This may be due to the effectiveness of tailored messaging employed in DrinksRation, with the literature recognizing its use for sustained adherence and successful outcomes^61,62^.

This study had several limitations. First, attrition at follow-up was substantial, which is a common challenge in digital health research and may reflect the difficulty of sustaining user engagement over time^63^. Although the primary analyses used a modified intention-to-treat repeated-measures approach that incorporated available outcome data under a missing-at-random assumption, loss to follow-up may still have introduced bias. Future studies should therefore explore strategies to improve retention, including adaptive or more dynamic intervention designs. A second challenge was the identification and management of fraudulent or suspicious participants^40^. Validation procedures, including IP address analysis, email validation, and manual checks, were important for maintaining data integrity. These issues are increasingly recognized in online research and necessitate ongoing attention and methodological improvements^40^. Finally, the generalizability of these findings to broader veteran or civilian populations remains unclear. While DrinksRation was tailored to female veterans, its efficacy and acceptability in broader demographics require further investigation.

As the use of online technologies such as smartphone apps and online surveys to conduct and collect data for research purposes increases, so has the percentage of fraudulent participants gaining access to surveys, creating concerns about the impact on data validity^64^. When examining respondents to their online survey, Silverstein and colleagues (2022^65^) determined that suspicious responses were most likely to come from bots identifying survey links with a financial incentive automatically on social media. They further found that bots were likely to use tools such as ChatGPT/Microsoft Copilot to complete open-text questions, giving superficially relevant or unconnected responses. We used several strategies to detect and minimize fraudulent participants from pre- to post-intervention. Where participant responses were considered suspicious, but we were unable to definitively discern the authenticity of the participant in question, they were closely monitored throughout the intervention period.

This study provides evidence that targeted digital interventions like DrinksRation effectively reduce alcohol consumption and hazardous drinking behaviours among female veterans, addressing an identified gap in service provision^4^. Given the recognition of female veterans as a previously under researched group, these results underscore the necessity for gender-specific digital health solutions capable of overcoming barriers to traditional healthcare access.

## Data Availability

Data and code syntax arising from the Randomized Controlled Trial is available via the Open Science Framework.

## Acknowledgments

We note with great sadness the passing of our co-author, Professor Nicola T. Fear, before the final version of this publication. Professor Fear made an important contribution to this work, and we are grateful for her scholarship, guidance, and enduring impact on the field.

## Notes

### Competing Interest Statement

The authors have declared no competing interest.

### Clinical Trial

NCT05970484

### Clinical Protocols

https://doi.org/10.2196/51531

### Funding Statement

Yes

### Author Declarations

This study was approved by the ethics committee of King’s College London (LRS/DP-22/23-36879).

